# Estimating the ascertainment rate of SARS-CoV-2 infection in Wuhan, China: implications for management of the global outbreak

**DOI:** 10.1101/2020.03.24.20042218

**Authors:** Theodore Lytras, Georgios Panagiotakopoulos, Sotirios Tsiodras

## Abstract

We sought to estimate the ascertainment ratio of SARS-CoV-2 infection in Wuhan, China, using a modified Bayesian SEIR model with publicly reported case data. We estimated it at 0.465% (95%CI: 0.464–0.466%), implying that the outbreak in Wuhan was abated by depletion of susceptibles, rather than public health action alone. This suggests a high-transmissibility/low-severity profile for the current pandemic and raises doubt about whether suppression, rather than mitigation, is a feasible goal.

---

The coronavirus disease 2019 (COVID-19) outbreak originated at the end of 2019 in the city of Wuhan, China, and is currently sweeping the globe, with more than 370,000 confirmed cases and 16,000 deaths. With the outbreak having already turned into the latest global pandemic, one of the key unknowns that will determine its public health impact is the Infection Fatality Rate (IFR), i.e. the number of deaths as a proportion of all persons infected with the SARS-CoV-2 novel coronavirus, the causative pathogen of COVID-19. The IFR is expected to be much lower than the Case Fatality Rate (CFR), the proportion among confirmed cases, if many SARS-CoV-2 infections with mild or no symptoms are not being ascertained [1]. A low ascertainment rate has been previously suggested, based on epidemic modelling of exported cases: between 1.8% and 14% in Wuhan [2–5], 28.4% in Italy [6] and just 0.23% in Iran [7].

After the lockdown that came into force on 23 January 2020, Wuhan approximates a closed population in which the outbreak appears to have run its course, with cases on a steady decline since mid-February. We sought to estimate the ascertainment rate by applying epidemic modelling to publicly reported confirmed COVID-19 cases in Wuhan; a secondary objective was to estimate other epidemic parameters, including the basic reproduction number R_0_.

## Estimating the ascertainment rate in Wuhan

We downloaded the most recent confirmed case data from Wuhan using the R package ‘nCov2019’ [8], for the period between 3 January and 9 March 2020. On 12 February there was a spike of 13,436 retroactively reported confirmed cases due to a change in the case definition; we addressed this by linearly interpolating the new confirmed cases for that date and distributing the remainder to all previous dates in proportion to the number of cases already reported.

The median incubation period of COVID-19 has been estimated at approximately 5-6 days, with a range between 1-14 days [9,10]. According to the WHO-China joint mission report, the median time from symptom onset to laboratory confirmation in Wuhan decreased from 15 days (range 10-21) in “early January” to 5 days (3-9) in “early February” [11]. To reflect the above, we modelled the time between case reporting and symptom onset with a Weibull distribution with a shape/scale parameter of 8/15.7 on January 10, decreasing linearly to 4/5.5 on February 10 and after. We similarly modelled the time between case reporting and case exposure (infection with SARS-CoV-2) with a Weibull distribution with shape/scale 4.4/22.3 decreasing linearly to 2.6/12.1 (Figure 1). These distributions were used to reconstruct estimated epidemic curves by infection date and by symptom onset, up until February 23, discarding the last two weeks available.

**Figure 1:**
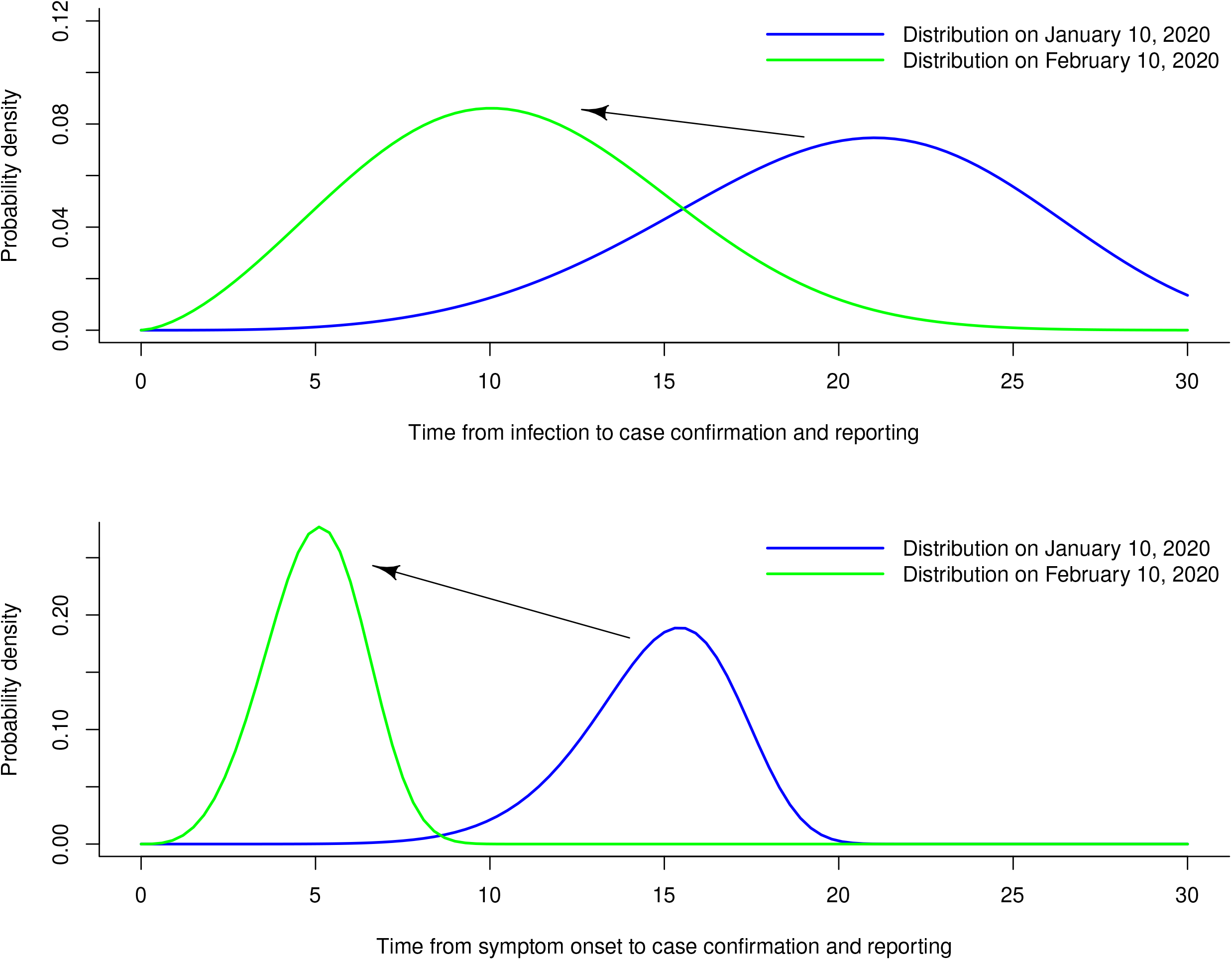
Assumed distributions for time between case infection and case reporting, and time between symptom onset and case reporting

To these data we subsequently fitted a discrete-time SEIR (Susceptible-Exposed-Infected-Removed) model, modified for underascertainment and fitted in a Bayesian framework. The model assumes that cases are observed from only a fraction of the population (i.e. the ascertainment rate) that is stable over time. We further assumed that cases are isolated upon confirmation and are thus removed from the “Infected” pool at that time. Transition probabilities at time *t* between S-E, E-I and I-R are 1-exp(-*β*\**I*_*t*-1_/*N*), 1-exp(-*κ*) and 1-exp(-*γ*) respectively, where *I*_*t*-1_ the total number of all infected at time *t*-1, *N* the population of Wuhan (11 million people), *β* the contact rate, *γ* the removal rate and *κ* the incubation rate (the inverse of the incubation period). In this model, *β*/*γ* equals the basic reproduction number *R*_*0*_ and (1/*κ* + 1/2*γ*) equals the serial interval [12]. We used non-informative Uniform(0,10) priors for *β, γ* and *κ*, and Uniform(0,1) for the ascertainment rate. Four sensitivity analyses were performed: (1) a model with a variable contact rate in three roughly equal periods, i.e. until the lockdown, until 10 February and until 23 February, (2) variable contact rate and an assumed 10% ascertainment rate, (3) assuming a 85% drop in contact rate after the lockdown, as recently suggested [13], and (4) a simplified SIR model (rather than SEIR). All analyses were done with R and JAGS, using 4 chains of 20000 iterations each and another 5000 as burn-in. Model estimates are presented as posterior medians and 95% Credible Intervals (CrI). The full analysis code is available on GitHub: https://github.com/thlytras/nCoVasc.

## Results

A total of 49,948 confirmed COVID-19 cases were reported in Wuhan until 9 March, of whom 46,159 were reported until February 23. Figure 2 illustrates the confirmed cases and the estimated epidemic curves by symptom onset and infection date. All model results for the main and sensitivity analyses are detailed on Table 1. The ascertainment rate was estimated at just 0.465% (95% CrI 0.464–0.466%), meaning that for every confirmed case in Wuhan there had been approximately 200 SARS-CoV-2 infections that were not detected. The R_0_ was estimated at 3.07 (95% CrI 3.02–3.11), and the incubation period and serial interval at 4.4 and 8.8 days respectively; the latter remained identical in sensitivity analyses.

**Table 1:** Model-based estimates for the COVID-19 epidemic parameters in Wuhan, China.

| Parameter | Estimate (95% CrI) |
| --- | --- |
| *Main analysis* |  |
| Ascertainment rate (%) | 0.465 (0.464–0.466) |
| Basic Reproduction Number ($R_0$) | 3.07 (3.02–3.11) |
| Incubation period (days) | 4.38 (4.34–4.41) |
| Serial interval (days) | 8.80 (8.75–8.87) |
| *Sensitivity analysis 1 (variable contact rate)* |  |
| Ascertainment rate (%) | 0.480 (0.477–0.484) |
| Basic Reproduction Number ($R_0$) (until lockdown) | 5.33 (5.23–5.42) |
| Basic Reproduction Number ($R_0$) (until Feb 10) | 2.09 (2.05–2.13) |
| Basic Reproduction Number ($R_0$) (until Feb 23) | 3.83 (3.62–4.04) |
| Incubation period (days) | 4.38 (4.34–4.42) |
| Serial interval (days) | 8.81 (8.75–8.87) |
| *Sensitivity analysis 2 (variable contact rate, and 10% ascertainment rate assumed)* |  |
| Basic Reproduction Number ($R_0$) (until lockdown) | 4.13 (4.06–4.20) |
| Basic Reproduction Number ($R_0$) (until Feb 10) | 0.85 (0.84–0.87) |
| Basic Reproduction Number ($R_0$) (until Feb 23) | 0.45 (0.44–0.46) |
| Incubation period (days) | 4.38 (4.34–4.41) |
| Serial interval (days) | 8.81 (8.75–8.87) |
| *Sensitivity analysis 3 (assuming 85% drop in contact rate after lockdown)* |  |
| Ascertainment rate (%) | 0.930 (0.899–0.965) |
| Basic Reproduction Number ($R_0$) (until lockdown) | 5.86 (5.76–5.97) |
| Basic Reproduction Number ($R_0$) (until Feb 10) | 0.88 (0.86–0.89) |
| Incubation period (days) | 4.38 (4.33–4.41) |
| Serial interval (days) | 8.81 (8.75–8.87) |
| *Sensitivity analysis 4 (simplified SIR model)* |  |
| Ascertainment rate (%) | 0.513 (0.510–0.516) |
| Basic Reproduction Number ($R_0$) | 2.29 (2.25–2.32) |
| Serial interval (days) | 13.87 (13.74–14.00) |

**Figure 2:**
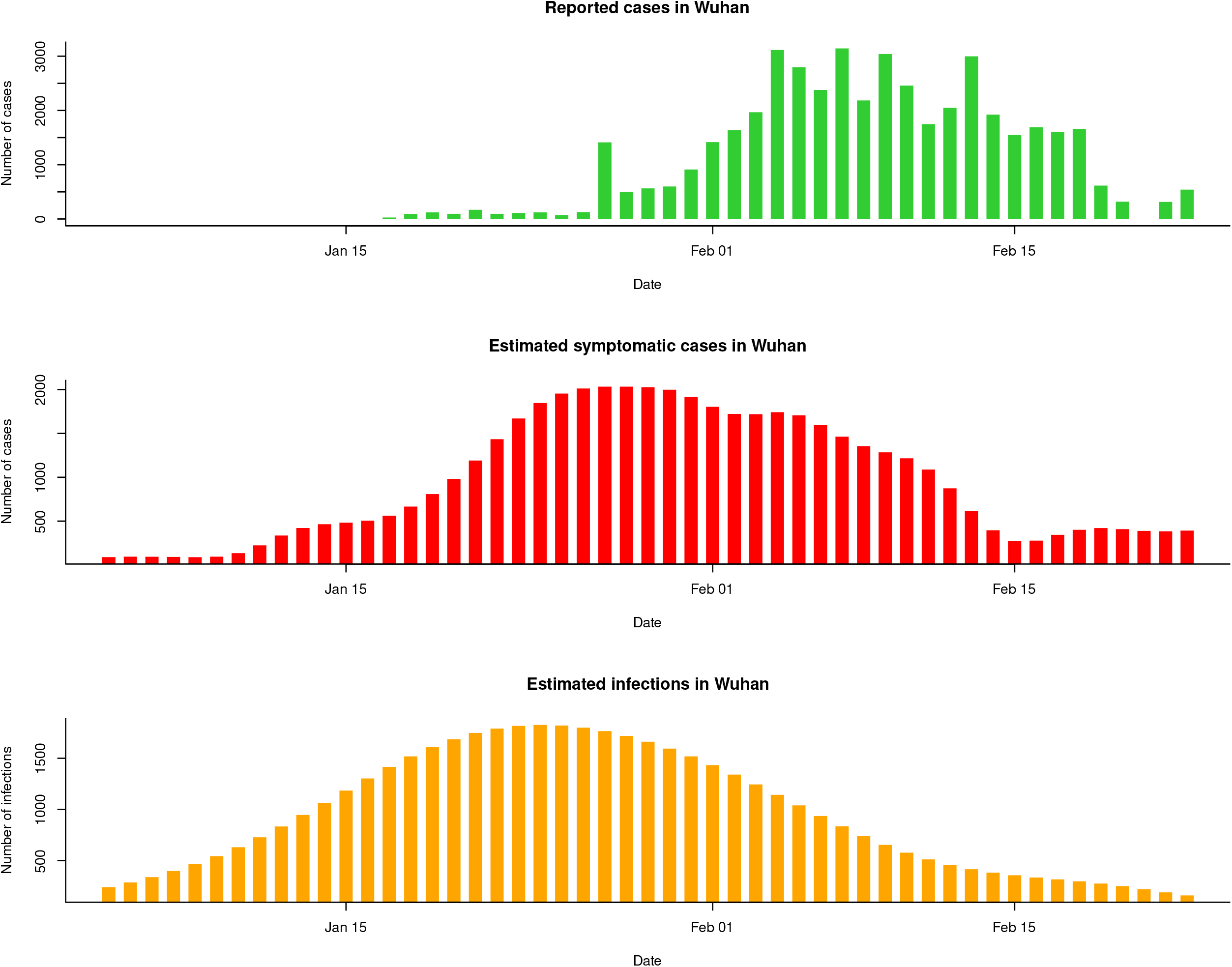
Cases of COVID-19 in Wuhan by reporting date, symptom onset (estimated) and time of infection (estimated)

Assuming a variable contact rate over the study period, the R_0_ estimate would be as high as 5.33 until the Wuhan lockdown, falling to 2.09 until 10 February then up again to 3.83, with an ascertainment rate of 0.48%. A nearly identical rate of 0.51% was obtained with a simplified SIR model, and a slightly higher rate of 0.93% assuming an 85% drop in contact rate after the lockdown. On the other hand, if one assumes a fixed ascertainment rate of 10% and a variable contact rate, the R_0_ would need to be as low as 0.85 right after the lockdown, and yet lower to 0.45 after 10 February, in order to fit the data (Table 1, sensitivity analysis 2).

The projected course of the epidemic in Wuhan, under the main analysis model, is shown in Figure 3. We estimate that half the population was already infected by 27 January, and by 23 February only 585,000 susceptibles remained (95% CrI 555,000–617,000), meaning 94.7% of the population had been infected. Assuming a 85% drop in contact rate after the lockdown (sensitivity analysis 3), the same percentage drops to 52.7% (95% CrI 51.0–54.4%).

**Figure 3:**
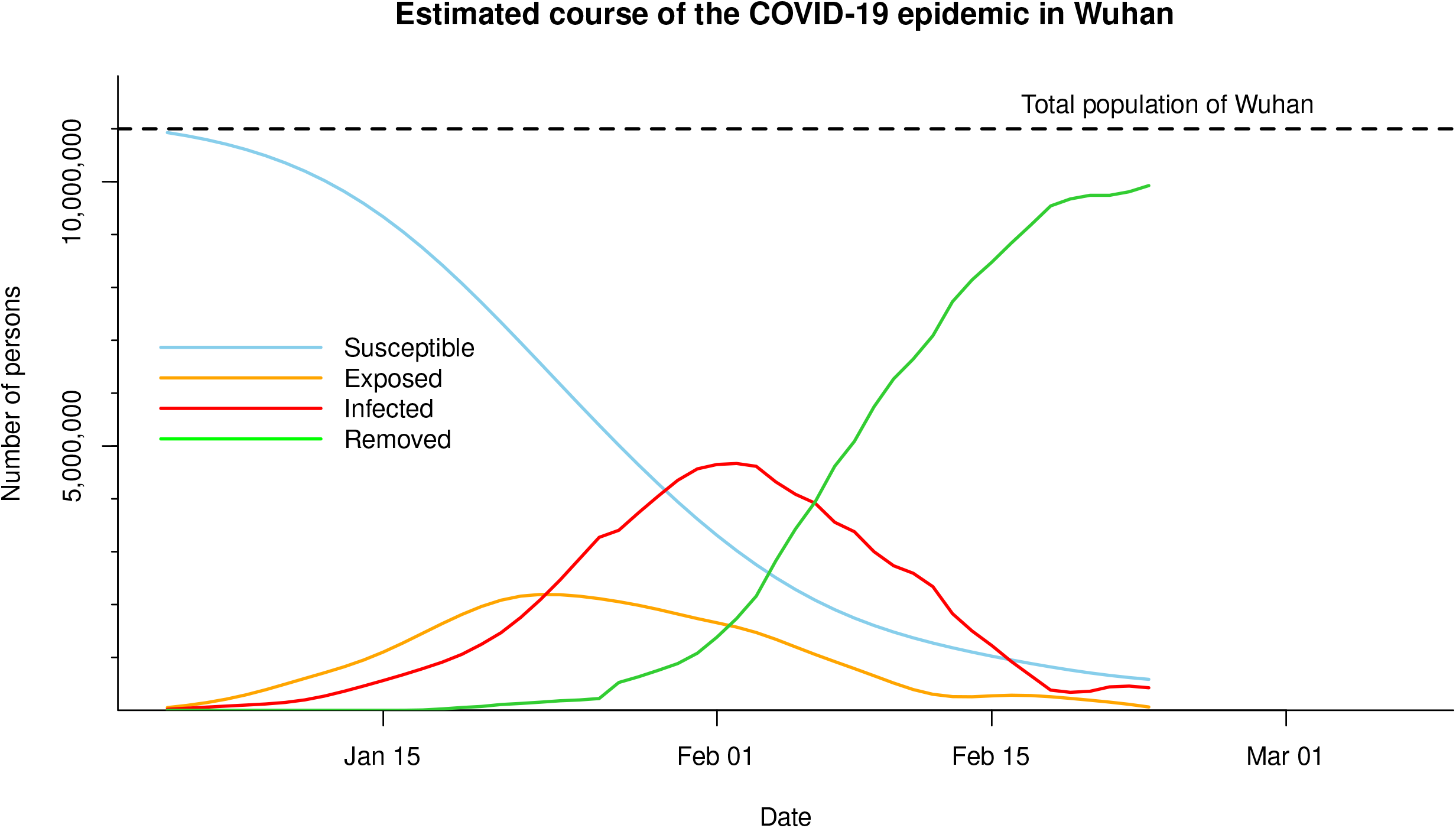
Estimated Susceptible, Exposed, Infected and Recovered population from COVID-19 over time in Wuhan

### Implications for the COVID-19 pandemic

It has been suggested that the COVID-19 outbreak in Wuhan was controlled via non-pharmaceutical measures and bold public health action that “changed the course of a rapidly escalating epidemic” [11]. Our modelling suggests another plausible scenario: that the large majority of the population was infected, with consequent depletion of susceptibles, and only a very small percentage of infections were ascertained. Otherwise, if ascertainment is assumed higher or not taken into account, very large reductions in transmissibility of SARS-CoV-2 are needed to account for the observed data, with about 80% of infections prevented immediately after the lockdown and 90% after February 10. It should be emphasized that in no way do our results diminish the value of non-pharmaceutical public health interventions in mitigating the outbreak, nor do they underestimate the immense efforts of the Chinese people. Their bold, large-scale action undoubtedly alleviated an already very difficult situation, and likely succeeded in “flattening the curve” of the pandemic to the maximum possible extent.

Accounting for case ascertainment is crucial for accurately assessing the true nature of the COVID-19 pandemic. An ascertainment rate as low as 0.5% in Wuhan suggests that the IFR of the SARS-CoV-2 virus is orders of magnitude lower than the CFR of COVID-19 (although some underascertainment of COVID-19 deaths should be taken into account as well). Together with a R_0_ estimate of 3.07, it would make COVID-19 the archetype of a high-transmissibility / low-severity pandemic, whose high impact is driven by the sheer number of infections rather than the virulence of the pathogen [14]. Our result further implies a very large amount of transmission from asymptomatic or minimally symptomatic persons, raising doubts about the feasibility of suppressing, much less containing, the COVID-19 pandemic [15]. Currently the World Health Organization (WHO) still advocates for a “containment strategy”, with laboratory testing for every suspected case, isolation of confirmed cases (including mild cases) in health facilities, and tracing all contacts [16]. Laboratory testing is indeed a priority for surveillance purposes, and also for severely ill patients, healthcare workers and other selected groups [17]. Given our findings, however, exhaustive testing and contact tracing are probably not optimal at this point in countries with ongoing epidemics, and risk detracting from the only eminently achievable end goal, which is mitigation: “flattening the curve” as much as possible, reducing stress on healthcare facilities and protecting the vulnerable. That is what is likely occurring in other locked-down areas of China right now, with the virus on a slow and stealthy progression, rather than on a freeze. Given the severity of the pandemic threat, no effort should be spared and no public health intervention discounted a priori. But at the same time, it is crucial to set realistic priorities and realistic expectations, as well as allocate resources optimally while advocating for more.

Our estimated ascertainment rate is on the lower end of previously published estimates for Wuhan [4]. Regardless of the precise rate though, the main conclusion about the feasibility of “containment/suppression” (or lack thereof) still holds, as also evidenced by the rapid worldwide spread of SARS-CoV-2. Our result also highlights the urgent need for seroprevalence surveys in different populations, to formally elucidate the true prevalence of the infection and guide further management of this new global pandemic. Striving for more timely data remains essential for optimal public health decisions.

## Data Availability

All data and code are publicly available on GitHub

https://github.com/thlytras/nCoVasc

## Acknowledgements

We gratefully acknowledge Dr. Vana Sypsa for providing very useful comments on this manuscript

